# Multiband EEG signature decoded using machine learning for predicting rTMS treatment response in major depression

**DOI:** 10.1101/2024.09.22.24314146

**Authors:** Alexander Arteaga, Xiaoyu Tong, Kanhao Zhao, Nancy B. Carlisle, Desmond J. Oathes, Gregory A. Fonzo, Corey J. Keller, Yu Zhang

**Author notes:** Corresponding author: Yu Zhang, PhD Assistant Professor of Bioengineering Assistant Professor of Electrical and Computer Engineering Lehigh University Bethlehem, PA 18015, USA.

## Abstract

Major depressive disorder (MDD) is a global health challenge with high prevalence. Further, many diagnosed with MDD are treatment resistant to traditional antidepressants. Repetitive transcranial magnetic stimulation (rTMS) offers promise as an alternative solution, but identifying objective biomarkers for predicting treatment response remains underexplored. Electroencephalographic (EEG) recordings are a cost-effective neuroimaging approach, but traditional EEG analysis methods often do not consider patient-specific variations and fail to capture complex neuronal dynamics. To address this, we propose a data-driven approach combining iterated masking empirical mode decomposition (itEMD) and sparse Bayesian learning (SBL). Our results demonstrated significant prediction of rTMS outcomes using this approach (Protocol 1: r=0.40, p<0.01; Protocol 2: r=0.26, p<0.05). From the decomposition, we obtained three key oscillations: IMF-Alpha, IMF-Beta, and the remaining residue. We also identified key spatial patterns associated with treatment outcomes for two rTMS protocols: for Protocol 1 (10Hz left DLPFC), important areas include the left frontal and parietal regions, while for Protocol 2 (1Hz right DLPFC), the left and frontal, left parietal regions are crucial. Additionally, our exploratory analysis found few significant correlations between oscillation specific predictive features and personality measures. This study highlights the potential of machine learning-driven EEG analysis for personalized MDD treatment prediction, offering a pathway for improved patient outcomes.

## Introduction

Major Depressive Disorder (MDD) is one of the most common mental illnesses worldwide, characterized by its symptoms of persistent sadness or emptiness, loss of appetite, trouble sleeping, and thoughts of suicide^1^. In 2020, it was estimated globally there were 3153 cases of MDD per 100000 population^2^. It can be severely debilitating; causing 49 million disability adjusted life years (years lost due to a disability) in 2020 alone^2^. The economic burden of MDD in the US has risen substantially with a 37.9% increase^3^, from $236.6 billion in 2010 to $326.2 billion in 2018. Failure to achieve a treatment response will lead to further harm and costs^4^. Current treatment strategies for moderate to severe MDD primarily include pharmacotherapy such as selective serotonin reuptake inhibitors ^44, 45, 46^. However, a considerable portion of patients are non-responders, known as treatment (medication) resistant ^47^. For these individuals, brain stimulation, particularly repetitive transcranial magnetic stimulation (rTMS), has emerged as an FDA-cleared and effective alternative treatment option for some ^5,6,7,8,25^. Further, there is a critical need to identify biomarkers for predicting treatment outcomes due to the biologically heterogeneous nature of MDD and the heterogeneity in rTMS treatment response, which often complicates treatment outcomes. Developing objective brain biomarkers (or signatures) is essential to reduce the need for multiple treatment trials and to expedite remission by more accurately selecting treatments.

Efforts to predict treatment outcomes and uncover the underlying neurophysiology in MDD patients have utilized neuroimaging techniques such as functional magnetic resonance imaging (fMRI) and electroencephalography (EEG) in combination with machine learning techniques^9,10,11,35^. While fMRI is an important and successful tool in neuroimaging, it is less clinically translatable due to high costs, the need for expertise, and specialized equipment. In contrast, EEG is more cost-effective, easier to deploy, and offers higher temporal resolution^36^.

Many studies utilizing machine learning and EEG in psychiatry have focused on EEG-based functional connectivity to identify promising biomarkers. While these studies have laid important groundwork, identifying treatment predictive biomarkers by decoding EEG spatio-temporal patterns at the electrode level represents an alternative approach^19,21^, especially at lower EEG channel densities. Despite these advances, very few studies have focused on identifying biomarkers for rTMS treatment using these methods. Additionally, many studies that have attempted to identify biomarkers for rTMS treatment have typically not examined multiband features jointly and are not end-to-end models^48^ (models that can learn to extract features directly relevant to the outcome), potentially resulting in lower prediction performance due to a loss of information. A modern method for end-to-end EEG decoding is the Sparse Bayesian Learning for End-to-End Spatio-Temporal-Filtering-Based Single-Trial (SBLEST) algorithm proposed by Wang et al^12^. This end-to-end sparse Bayesian learning framework aims to integrate spatial and multiband information to better understand channel-level features related to a specific EEG decoding task.

Conventionally, channel-level disorder-specific features are extracted by bandpass filtering a signal to a frequency band, typically one of the five where neural oscillations occur: delta (0.5-4 Hz), theta (4-8 Hz), alpha (8-13 Hz), beta (13-30 Hz), and gamma (30-100 Hz)^31^. However, distinct neuronal patterns may exist within the same band, potentially grouping multiple oscillations together, and oscillations between bands may be unintentionally attenuated^9^. Huang et al.^14^ introduced Empirical Mode Decomposition (EMD), a data-driven approach that decomposes a complex signal into simpler components without assuming it falls within a specific bandwidth or is sinusoidal, enabling detailed temporal and spectral feature extraction of nonstationary EEG signals. Despite its utility, EMD suffers from mode mixing in complex signals with low signal-to-noise ratios^15^. Spatial filtering in EEG decoding, using algorithms such as common spatial patterns (CSP), constructs spatial filters to isolate brain activity from specific regions, improving the signal-to-noise ratio^37^. These are typically employed before input into a machine learning model. In contrast, SBLEST learns spatial filters within its linear regression model, optimizing them for specific tasks while being computationally efficient.

In this paper, we are thus motivated to use a recent improvement of EMD, iterated masking empirical mode decomposition (itEMD), introduced by Fabus et al^16^, in combination with SBLEST to extract both spatial and temporal features in a data driven manner. The itEMD method is a completely data driven way to decompose signals into components with significantly less mode mixing^16^. SBLEST learns spatial filters within its linear regression model, optimizing them for specific tasks while being computationally efficient. Thus, we propose a combined methodology, EMD-SBL, to predict treatment outcome of MDD patients receiving rTMS using patient EEG data from the TDBRAIN dataset^17^. By utilizing the EMD-SBL framework, we aim to identify channel-level spatial features associated with chosen decomposed components. Furthermore, we examine the association between predictions from each signal component and NEO Five-Factor Inventory-3 personality scores (NEO-FFI) clusters to identify personality traits correlated to each channel-level EEG topographic map. MDD treatment response has been found to correlate with some NEO-FFI clusters for medication response^33^ but the opposite has been true rTMS response^42^. EMD-SBL provides unique insights by integrating spatial and temporal information through data-driven decomposition and optimized spatial filtering, offering a more nuanced understanding of EEG patterns and their relationship with treatment outcome in MDD patients receiving rTMS treatment.

## Results

### EMD-SBL predicts rTMS treatment response for MDD patients

For both rTMS protocols, we sought to create a model that could predict treatment response. Significant results were only achieved with eyes-open rsEEG data (See eyes-closed results in Supplementary Table 3). For Protocol 1 (N=44), we achieved the best performance using the EMD-SBL method (Pearson’s r = 0.401, p = 0.00703, p_permute_ < 0.05). Likewise, for Protocol 2 (N=73), the best performance is achieved using the EMD-SBL method (Pearson’s r = 0.255, p = 0.0292, p_permute_ < 0.05). Figure 2 displays the results of EMD-SBL for Protocol 1 and Protocol 2. Only our method achieved significant results when compared against the results for SBLEST + Band Filtering, SBLEST, and Ridge Regression + itEMD (Figure 3, Supplementary Tables 1 and 2).

**Figure 1.**
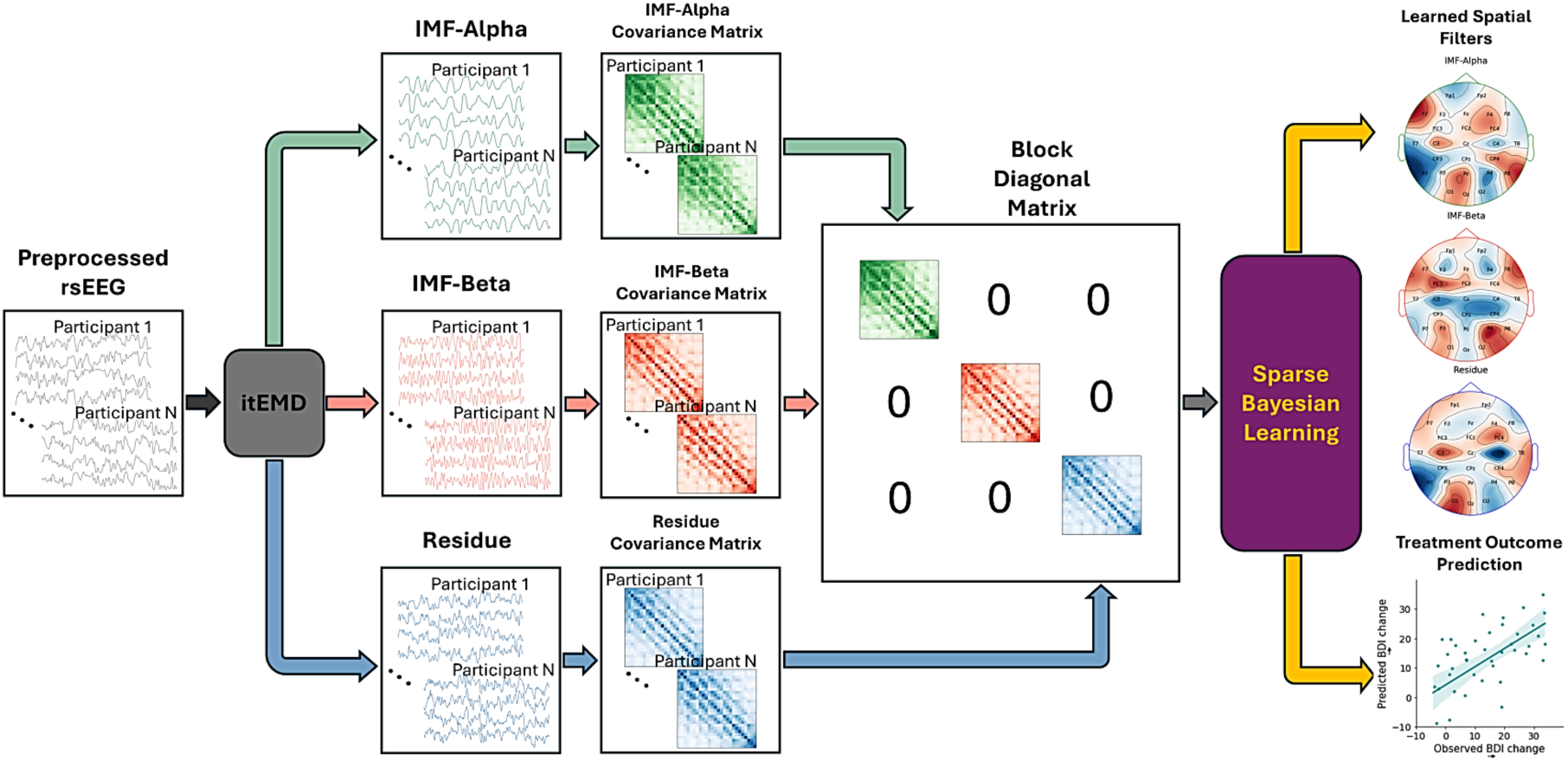
Illustration of the proposed EMD-SBL model for treatment outcome prediction and brain signature identification. The preprocessed EEG data was first decomposed using itEMD into multiple oscillation components: IMF-Beta, and IMF-Alpha, Residue (Original signal with IMF-Beta and IMF-Alpha subtracted). For each component, we calculated the covariance matrix and enhanced it using trace normalization, whitening, and logarithm transform. These matrices are combined to form a block diagonal matrix, which serves as the input for the SBLEST model.

**Figure 2.**
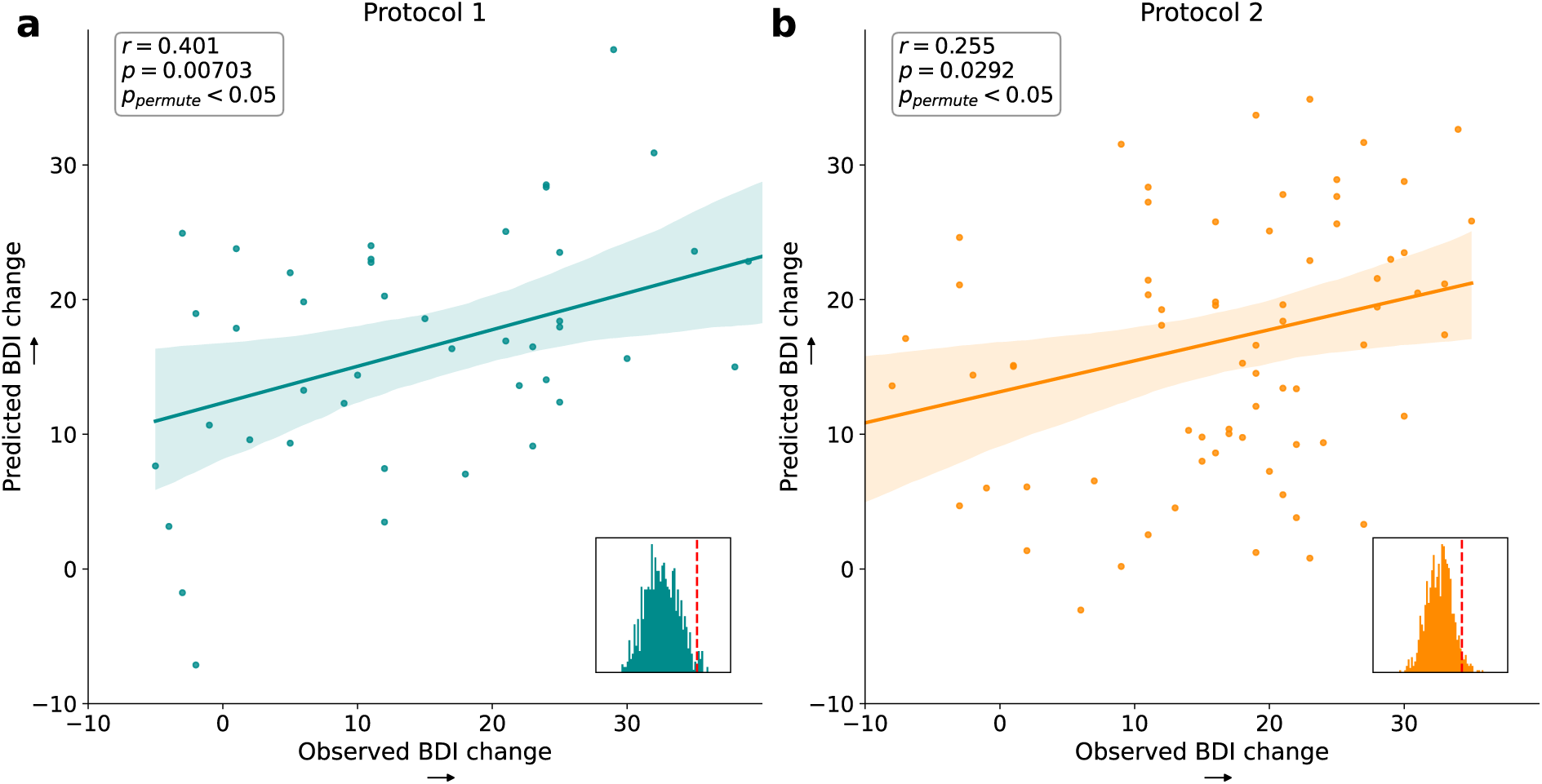
Prediction results of EMD-SBL model. The prediction was evaluated using 10 repetitions of 10-fold cross-validation for each protocol separately. Permutation test was done with 1000 runs **a.** Prediction of BDI change for Protocol 1 (N=44, Pearson’s r = 0.401, p = 0.00703, p_permute_ < 0.05) **b.** Prediction of BDI change for Protocol 2 (N=73, Pearson’s r = 0.255, p = 0.0292, p_permute_ < 0.05).

**Figure 3.**
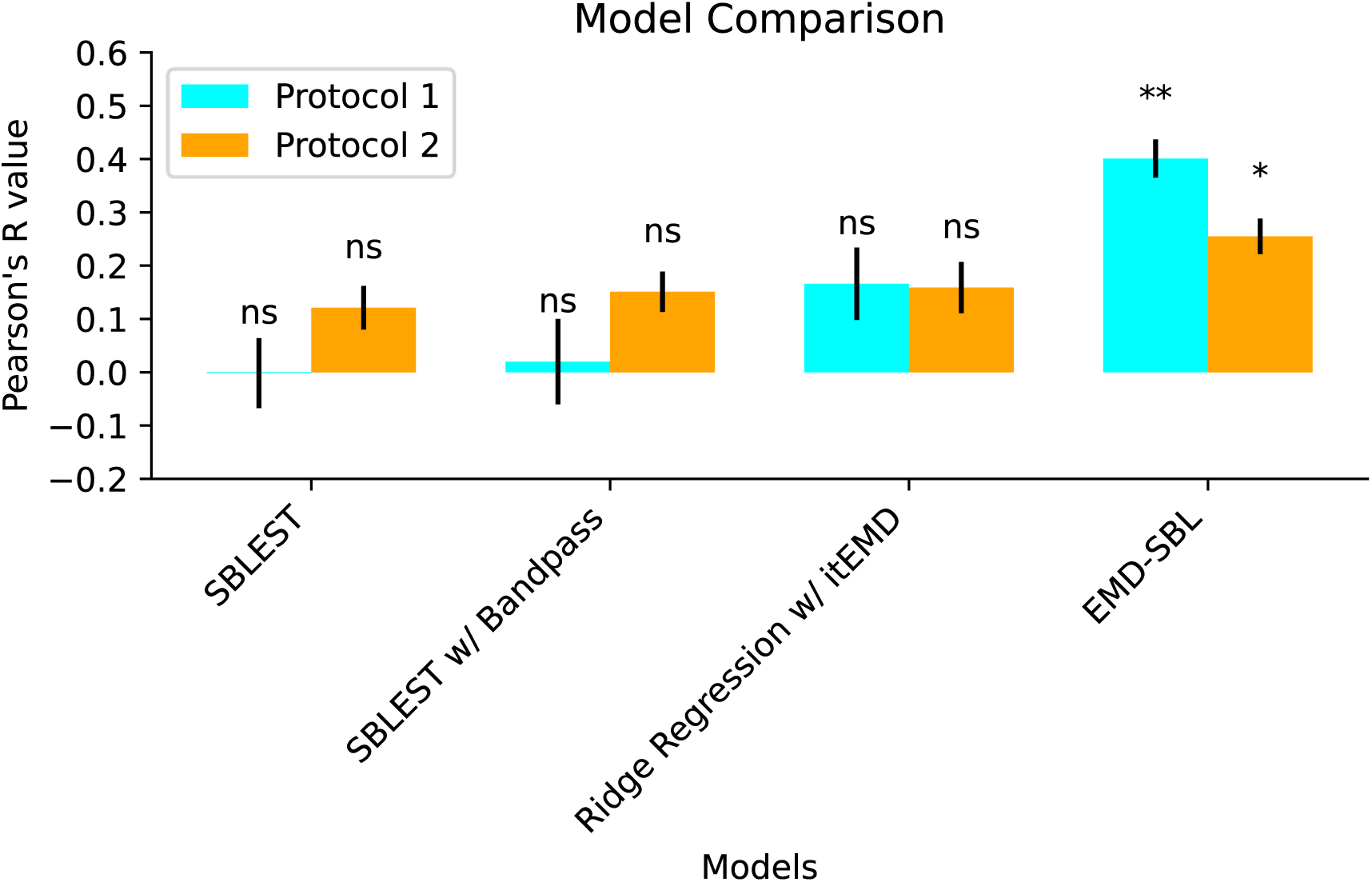
Model comparison results for SBLEST, SBLEST + Band Filtering, Ridge Regression + itEMD, and EMD-SBL. Only EMD-SBL was significant (** : p<0.01, * : p<0.05, ns : not significant).

### Treatment-predictive Signatures Interpretation

Next, we sought to interpret the identified biomarkers by examining model weights and spatial filters. We examined stability between runs by measuring the similarity of their weight matrix *W*. *W* is the weight matrix optimized by the SBL model. To obtain our treatment outcome prediction, we perform matrix multiplication between *W* and the block matrix of the test split. to Our model training incorporated three different oscillations (i.e. IMF-Alpha, IMF-Beta, and Residue components), allowing *W* to be split into components for oscillation-specific spatial pattern interpretation. **Figure 3** illustrates the stability and optimized matrices of each oscillation for each rTMS protocol. Following the method described by Wang et al^12^, we decomposed the learned symmetric matrix *W* into spatial filters and their regression weights. Through eigenvalue decomposition, we obtained leading eigenvalues (regression weights) and eigenvectors (spatial filters). The eigenvectors represent spatial filters, which we visualized over a 2D scalp map to better interpret the oscillations. **Figure 4** shows the topographical 2D channel mappings for the spatial filters of each oscillation. For Protocol 1 (10Hz L-DLPFC), IMF-Alpha had the highest magnitude eigenvalue, followed by IMF-Beta and then the Residue. Examining the spatial filters for each oscillation, we identified the channels with the highest absolute values in terms of importance for predicting treatment outcomes. The most important channels were: F7 and P7 for IMF-Alpha; FC3, C3, and P4 for IMF-Beta; and C3, C4, P7, and O1 for the Residue. In Protocol 2 (1Hz R-DLPFC), IMF-Alpha again had the highest magnitude eigenvalue, followed by the Residue and then IMF-Beta. The channels with the most importance for Protocol 2 were: F7, F3, and P7 for IMF-Alpha; FC3 for IMF-Beta; and F4, FC3, Cz, P7, and O1 for the Residue.

**Figure 4.**
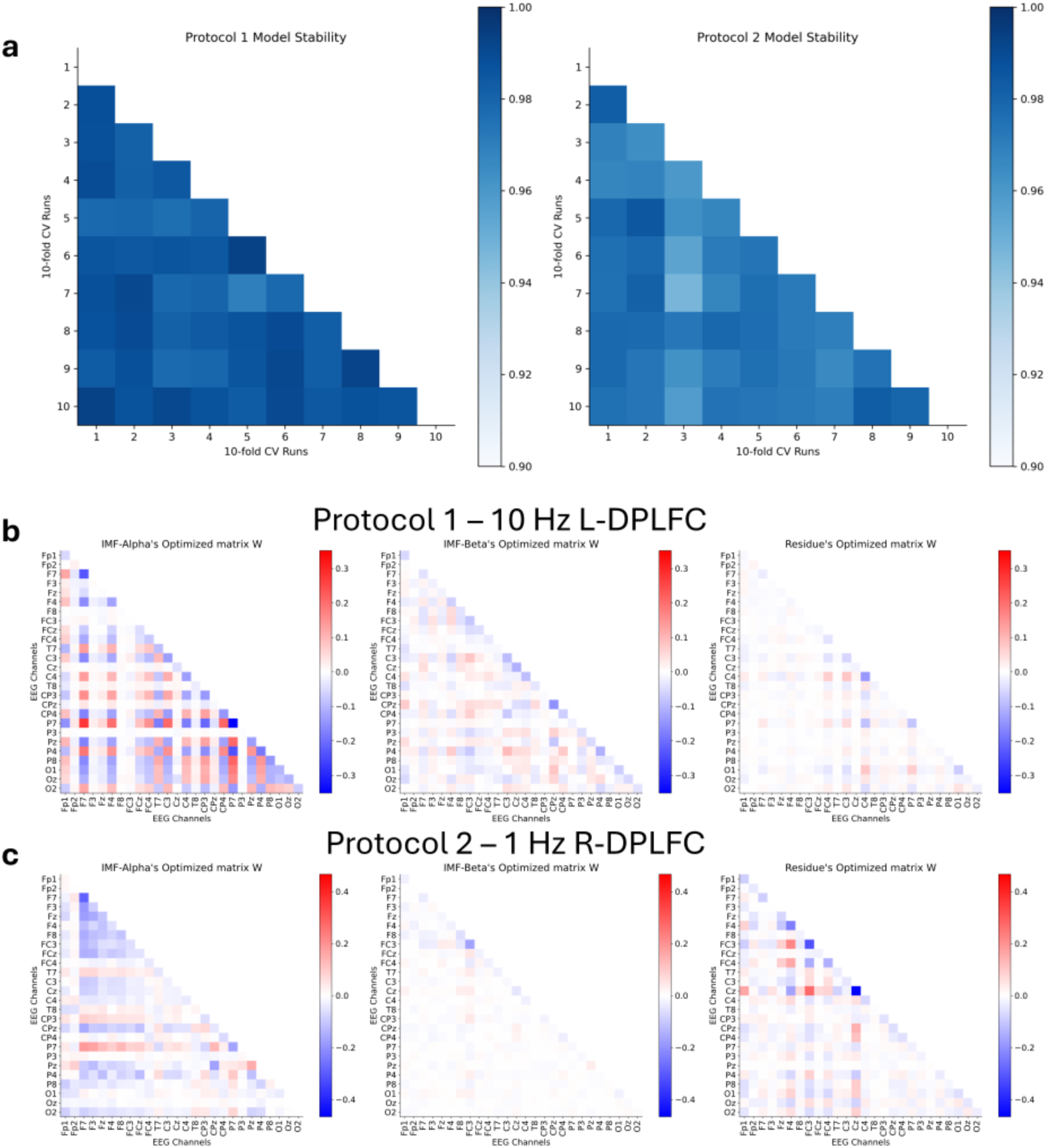
Learned model weights. **a.** Model stability across runs from 10 repetitions of 10-fold cross-validation. Stability was calculated by measuring the correlation coefficient of the learned matrix *W* between runs. **b.** Protocol 1 *W* matrix of each oscillation optimized by SBLEST. Between channel covariance, IMF-Alpha had the most channel feature importance. **c.** Protocol 2 *W* matrix of each oscillation optimized by SBLEST. Similar to Protocol 1, IMF-Alpha had the most channel feature importance.

### Oscillation NEO-FFI correlations

To further interpret the behavioral relevance of the identified biomarkers, we calculated the correlation coefficient between the oscillation’s predictions and the NEO-FFI items of each participant (shown in **Figure 5** and **Figure 6** for Protocols 1 and 2 respectively). For the participants in Protocol 1, the Residue had significant correlation (p<0.05) with self-approach, a neuroticism subscale, and orderliness, a conscientiousness subscale. Further, the predictive features of IMF-Beta were correlated with aesthetic interest, a subscale of openness. Of predictive features of Protocol 2, the Residue had a significant correlation with extraversion, openness and its subscale intellectual interest, and conscientiousness.

**Figure 5.**
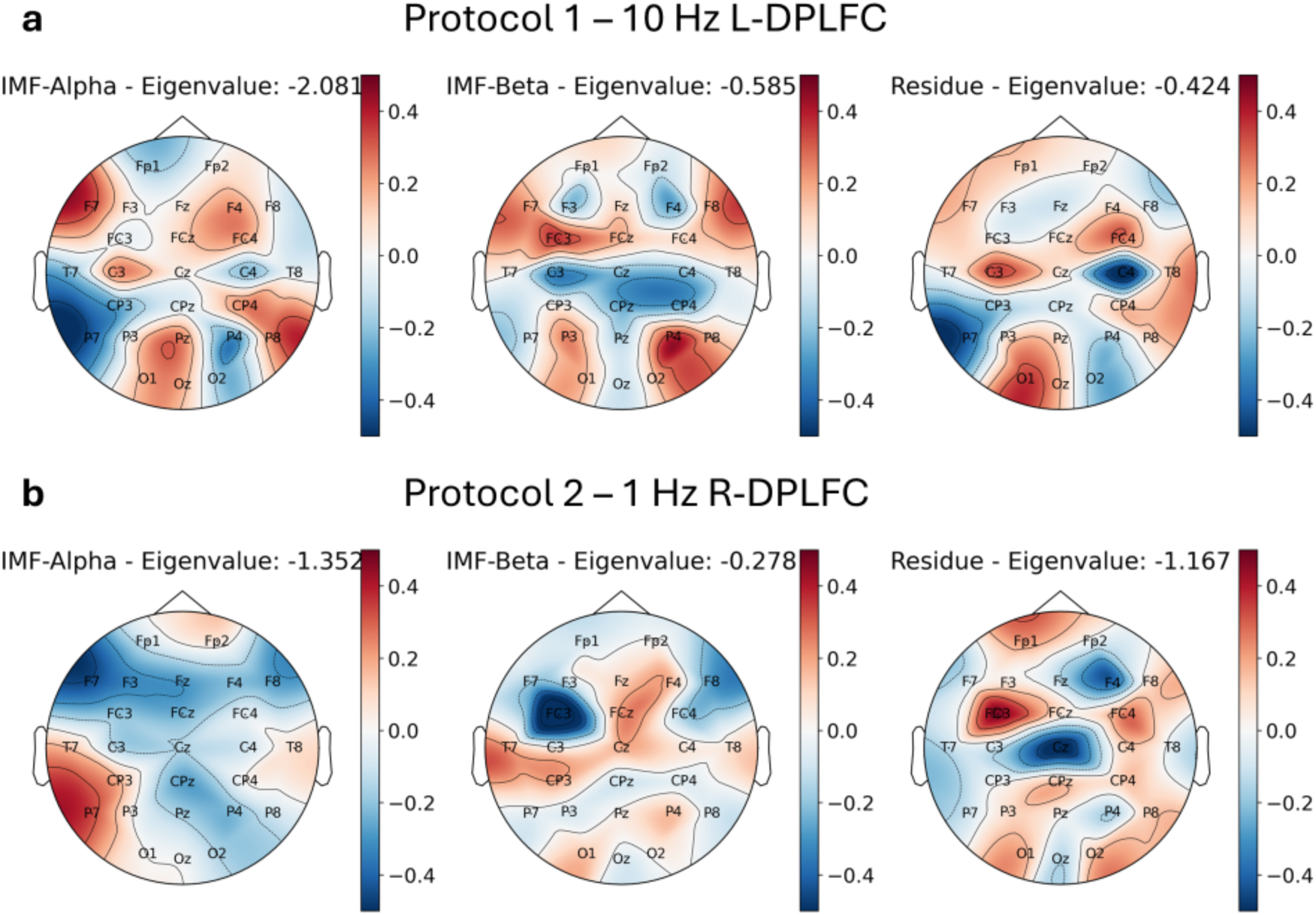
Spatial filter patterns. **a.** Spatial filters with their corresponding regression weight (eigenvalues) for Protocol 1 – 10Hz L-DLPFC. For IMF-Alpha, the channels with the most feature importance: F7, P7. For IMF-Beta: FC3, C3, P4. For the Residue: C3, C4, P7, O1. **b.** Spatial filters with their corresponding regression weight for Protocol 2 – 1Hz R-DLPFC. For IMF-Alpha the channels with the most feature importance: F7, F3, P7. For IMF-Beta: FC3. For the Residue: F4, FC3, Cz, P7, O1.

**Figure 6.**
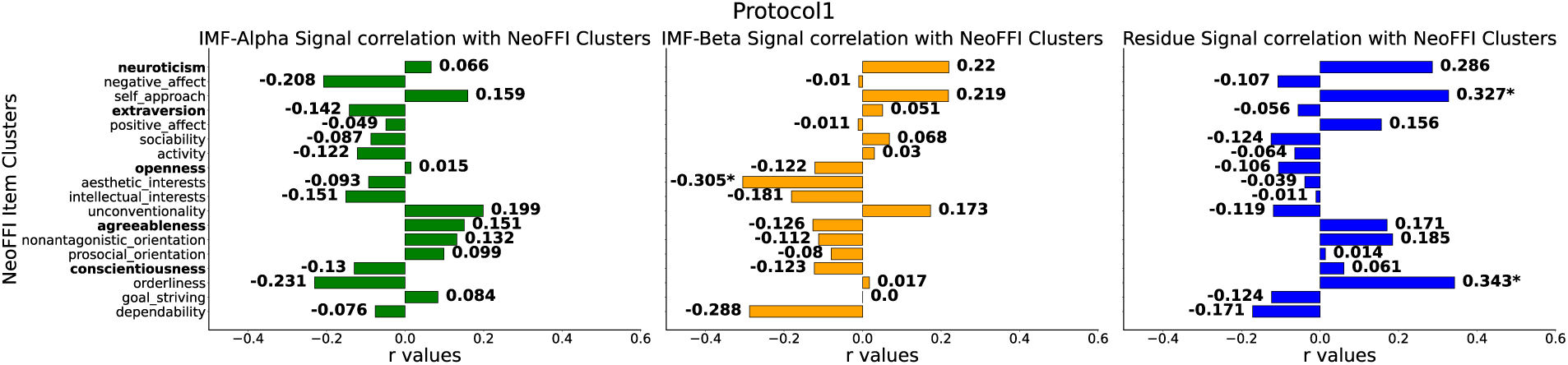
Association between the Protocol 1-predictive signature and NEO-FFI. **a.** Correlations for IMF-Alpha: No significant (p<0.05) correlations. **b.** Correlations for IMF-Beta: Significant (p<0.05) correlations with aesthetic interest (Pearson’s r = -0.305), a subscale of openness. **c.** Correlations for the Residue: Significant (p<0.05) correlations with self-approach (Pearson’s r = 0.327), a neuroticism subscale, and orderliness (Pearson’s r = 0.343), a conscientiousness subscale.

**Figure 7.**
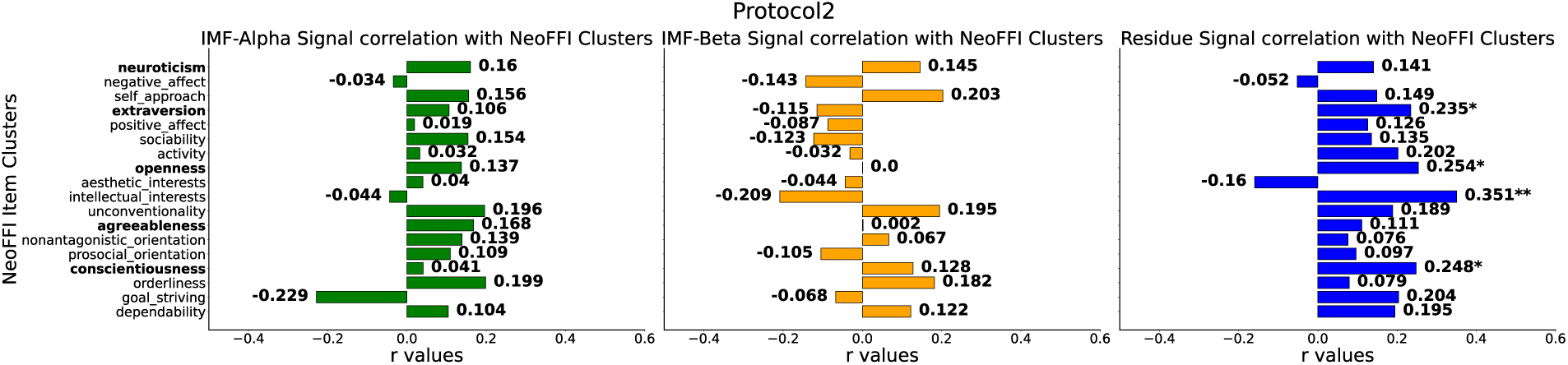
Association between the Protocol 2-predictive signature and NEO-FFI. **a.** Correlations for IMF-Alpha: No significant (p<0.05) correlations. **b.** Correlations for IMF-Beta: No significant (p<0.05) correlations. **c.** Correlations for the Residue: Significant (p<0.05) correlations with extraversion (Pearson’s r = 0.235), openness (Pearson’s r = 0.254) and its subscale intellectual interest (Pearson’s r = 0.351), and conscientiousness (Pearson’s r = 0.248).

## Discussion

In this study, we presented a novel methodology for revealing multimodal EEG signatures to predict rTMS treatment outcome for MDD patients. By combining itEMD and SBLEST, our EMD-SBL method captured more variability in treatment outcome compared to traditional bandpass filtering-based approaches. Traditional methods may lose clinically relevant information by assuming that neuro-oscillations are consistent across participants within the same frequency bands. EMD-SBL, in contrast, demonstrated significant prediction accuracy (p<0.05) for rTMS treatment outcome in both Protocols, highlighting its effectiveness. While our models for both Protocols demonstrated significant predictions, the model for Protocol 2 (1Hz R-DLPFC, N=73) performed worse than Protocol 1 (10Hz L-DLPFC, N=44). This discrepancy may be because IMF-Alpha is not as predictive for Protocol 2 and instead including a lower frequency IMF may have been more appropriate for Protocol 2. As shown in **Figure 3**, feature weights for IMF-Beta for Protocol 2 are negligible, whereas the Residue, derived by subtracting IMF-Beta and IMF-Alpha from the original signal, shows much higher feature importance. This suggests that other oscillations in the rsEEG may better predict treatment outcome for Protocol 2.

IMF-Alpha, closely associated with the alpha band, proved to be the most important oscillation for predicting treatment outcome in both protocols. This finding aligns with previous literature, which has identified alpha oscillations as highly predictive of treatment outcome in MDD treatments^27,32,35^. Other studies have also found a link between the alpha band frequency and treatment response in TMS. Conca et al. ^41^ found that initial lower mean alpha frequency of EEG was a possible predictor of TMS response as an add-on therapy. Arns et al. ^42^ found that a slower anterior individual alpha peak frequency was one of the differentiating factors between responders and non-responders in rTMS. In our dataset, participants received concurrent rTMS and psychotherapy for both protocols^25^. Therefore, the shared oscillation could also be predictive of psychotherapy response. Spatially F7 and P7 proved to have the highest importance in the alpha oscillations in both models, consistent with previous findings indicating alpha oscillations in the frontal and parietal areas as predictive of treatment response^38,39,40^.

IMF-Beta, similar to the beta band, was only predictive of Protocol 1. This oscillation may capture features specific to 10 Hz rTMS at the L-DLPFC. Previous research has indicated that beta oscillatory events in pretreatment EEG could indicate treatment response^28,43^. Additionally, we found that channels FC3, C3, and P4 contributed most to IMF-Beta for predicting treatment outcome, supporting the observation that frontal beta can predict rTMS treatment outcomes^28^.

Our further exploratory analysis correlated the predictive signature of each oscillation with NEO-FFI personality scores. Previous literature^33^ had indicated that traits such as neuroticism, extraversion and conscientiousness are associated with treatment outcomes in MDD patients who were treated with combined antidepressant-psychotherapy. However, the same could not be said for rTMS treatments, where no significant correlation between rTMS treatment response and the NEO-FFI items were found^42^. Our study aligns with this as our most informative features, IMF-Alpha and IMF-Beta, had no correlation with personality scores.

In summary, our combined itEMD and SBLEST methodology (EMD-SBL) showed promise in predicting rTMS treatment outcomes in MDD using rsEEG data from the TDBRAIN dataset. Our data-driven approach revealed critical brain signatures by jointly modeling multiband spatio-temporal filters from multichannel EEG. Many of the signatures are further validated by their appearance in existing literature. Our work advances the potential for treatment response prediction in rTMS, moving towards creating generalizable, cost-effective frameworks for more personalized treatments and better outcomes for individuals with MDD.

## Materials and Methods

### Participants

Participant data and demographics were obtained from the TDBRAIN dataset^17^. Only participants who were diagnosed with MDD and received rTMS were included (N=117) from the naturalistic open label study. Participants received either of the two rTMS protocols: high frequency rTMS (10Hz) at the left dorsolateral prefrontal cortex (DLPFC) (Protocol 1) or low frequency rTMS (1 Hz) at the right DLPFC (Protocol 2). For Protocol 1, 1500 pulses were administered at each rTMS session, while 1200 pulses were administered for Protocol 2. Sessions occurred two to three times a week. Depression severity was measured using the Beck Depression Inventory (BDI)^18^. If no response was observed by sessions 20–25, treatment is discontinued. If the BDI score indicated remission (defined as score ≤12) for five consecutive sessions, the patient was given the option to end treatment, phase out sessions by gradually reducing their frequency, or extend with maintenance sessions occurring once every 6–8 weeks. The primary treatment outcome was measured by pre-minus-post-treatment change in BDI. It is important to note that the original study found no significant difference in treatment outcome between these two protocols^25^. The NEO-FFI was used to assess the personality traits of the participants.

### EEG Recordings

Both eyes-open and eyes-closed resting state EEG (rsEEG) data was recorded from all the selected participants. Only the pre-treatment rsEEG was used. Data was collected from 26 channels of EEG for two minutes at a sampling rate of 500 Hz using a Neuroscan NuAmps amplifier. The electrode labels, based on the 10-10 electrode international system, are as follows: Fp1, Fp2, F7, F3, Fz, F4, F8, FC3, FCz, FC4, T7, C3, Cz, C4, T8, CP3, CPz, CP4, P7, P3, Pz, P4, P8, O1, Oz, O2.

### EEG Preprocessing

All the EEG data was preprocessed using an EEGLAB-based pipeline^19^. The preprocessing steps are as follows. The data was downsampled from 500 Hz to 250 Hz, followed by removal of 50 Hz line noise using notch filtering^20^. Low frequency noise near DC was removed from the data using a 0.01-Hz high-pass filter. Bad segments were rejected by thresholding the magnitude of each segment. Bad channels were detected by thresholding the spatial correlations among the 26 EEG channels and interpolated via the spherical spline interpolation^22^. Any individuals with more than 20% of channels detected as bad were excluded from our analysis. Independent component analysis (ICA) was further applied to remove artifacts such as eye blinks, muscular activity, and cardiac activity (ECG)^23^. Lastly, the EEG channels are re-referenced to the common average.

### Iterated Masked Empirical Mode Decomposition

In this study, we seek to utilize a multiband approach to fully leverage oscillation features in EEG data. Traditional bandpass filtering divides the data into canonical neural oscillation frequency bands, assuming uniform oscillation frequencies across patients. In contrast, EMD allows us to gain more detailed insights by decomposing the signal into intrinsic mode functions (IMFs). This method may help identify more effective biomarkers for predicting treatment outcomes by capturing patient-specific oscillatory modes. Each decomposed IMF represents a narrowband oscillatory mode with distinct frequency and amplitude characteristics. EMD decomposes a signal into a set of IMFs through an iterative process: 1) Identify the local extrema for one channel *X*(*t*). 2) Fit the maxima and minima to an envelope. 3) Calculate the mean of the upper and lower envelopes. 4) Subtract this mean from the original signal to obtain signal *H*(*t*). 5) Check if *H*(*t*) satisfies the conditions for an IMF. If yes, subtract the IMF from *X*(*t*) and repeat steps 1-5 with the resulting signal. If not, repeat steps 1-5 with *H*(*t*). 6) Continue this cycle until the residual signal no longer contains significant oscillations.

A limitation of traditional EMD is mode mixing, where one frequency component can appear in multiple IMFs^15^. To address this, we used masked EMD, which involves adding a masked signal to the original signal *X*(*t*), in step 1. By iterating this process with different masking signals, we obtain distinct IMFs. Iterated masked empirical mode decomposition (itEMD) further improves the masking technique by determining masked frequencies specific to the data, enabling a purely data-driven decomposition of the EEG signal. We follow the steps laid out by Fabus (2021)^16^: 1) Select an initial set of mask frequencies based on the dyadic masking technique. 2) Perform masked EMD to obtain IMFs. 3) Use the Hilbert Transform to calculate the Instantaneous Frequency (IF) for each IMF. 4) Calculate the amplitude-weighted average of each IF and set it as the next masked frequency. 5) Repeat steps 2-4 until the relative difference between the current and previous masked frequencies is minimal.

### EMD-SBL Multiband Prediction

Conventionally, decoding EEG spatial features involves algorithms such as CSP, which create spatial filters to maximize variance between classes and isolate brain activity from specific regions. Although this method enhances the signal-to-noise ratio for decoding task-relevant EEG patterns, it is limited by its reliance on assumptions about EEG sources and scalp potential distributions, which are not fully data-driven.

SBLEST is a novel EEG decoding algorithm employing a low-rank weight matrix within a sparse Bayesian learning framework^12^, which simultaneously optimizes spatio-temporal filters and prediction models. The SBLEST prediction model can be represented by the equation: *y_m_* = *tr*[*WR_m_*] + ε, where *W* is a symmetric weight matrix decomposable into spatial filters and regression weights, and *R_m_* is the covariance matrix of the EEG data and enhanced by trace normalization, whitening, and logarithm transform. The optimal model hyperparameters can be automatically estimated under a Bayesian framework based on all available training data without the need for cross-validation. This algorithm has shown superior performance, compared to contemporary methods, including deep learning approaches, on revealing meaningful neurophysiological patterns in five motor imagery datasets and one emotion recognition dataset^12^. Thus, we chose this model to predict treatment outcome and identify the underlying brain activity associated with the outcome. To leverage multiple EEG oscillations after itEMD decomposition, we modified SBLEST (**Figure 1**) to jointly learn spatio-temporal patterns from various oscillations, creating a multiband brain signature for robustly predicting rTMS treatment outcome. We constructed a block diagonal matrix using the enhanced (trace normalization, whitening, and logarithm transform) covariance matrices of different oscillations: the oscillation closest to beta (IMF-Beta), the oscillation closest to alpha (IMF-Alpha), and the residual oscillation after subtracting both oscillations from the original signals. We focused on IMF-Beta and IMF-Alpha based on previous literature showing statistical correlations between treatment outcome and alpha and beta oscillations^26,27,28,29,35^. The residue accounts for any remaining oscillations or signal components that might also be important in treatment outcome prediction. This approach assumes that the alpha, beta, and residual oscillations are independent of each other. The weight matrix *W* learned by the SBLEST model can be decomposed to generate spatial maps for different oscillations, providing a comprehensive view of the brain activity associated with treatment outcome.

### Model Evaluation

To reliably evaluate the machine learning models, we performed 10 repetitions of 10-fold cross-validation. During training, the training set was further augmented using the time reversal strategy^30^. The prediction model was evaluated by first taking the mean of the predicted value for each subject across all repetitions of the cross-validation. Then, we proceeded by calculating the Pearson’s correlation coefficient (r) and r-squared value (R^2^) between the predicted pre-minus post-treatment BDI change and the actual change.

## Data Availability

The TDBRAIN data is publicly available through: https://brainclinics.com/resources/.

## Acknowledgements

This work was supported by NIH grant nos. R01MH129694, R21MH130956, R21AG080425, Alzheimer’s Association Grant (AARG-22-972541), and Lehigh University FIG (FIGAWD35), CORE, and Accelerator grants. Portions of this research were conducted on Lehigh University’s Research Computing infrastructure partially supported by NSF Award 2019035. G.A.F. was also supported by philanthropic funding and NIH grant nos. R01MH132784 and R01MH125886, and grants from the One Mind - Baszucki Brain Research Fund, the SEAL Future Foundation, and the Brain and Behavior Research Foundation.

## Supplementary Information

**Supplementary Figure 1.**
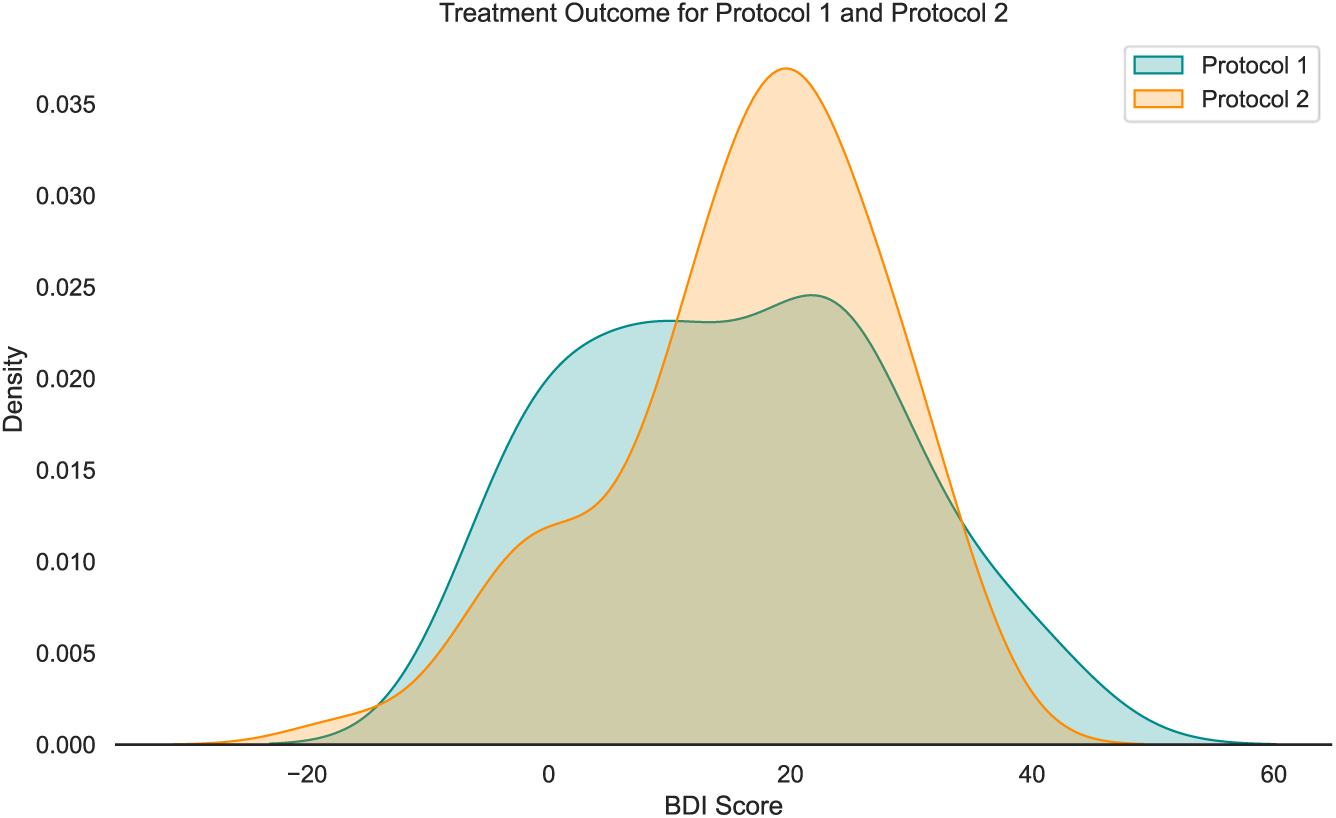
In blue, BDI score change distribution after patients with MDD received 10 Hz rTMS at the left DLPFC (Protocol 1 N=44). In orange, BDI score change distribution after patients with MDD received 1 Hz rTMS at the right DLPFC (Protocol 2 N=73).

**Supplementary Figure 2.**
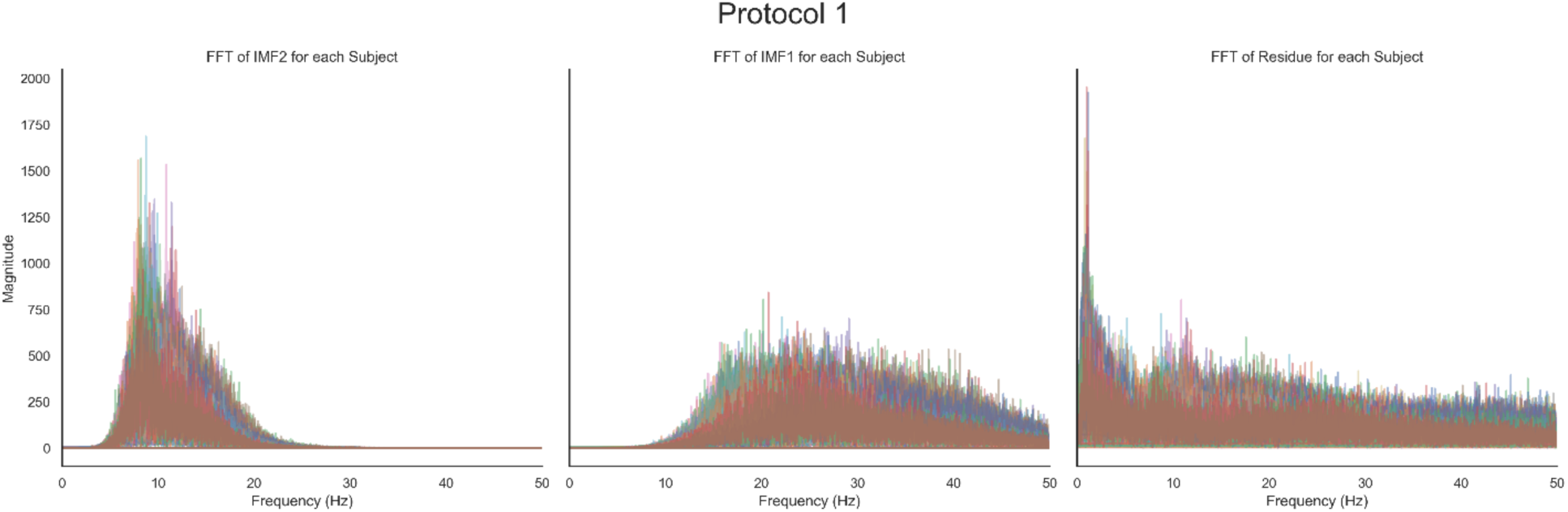
FFT of IMF2, IMF1, and the Residue for Protocol 1 at electrode F3 for each subject. From visual inspection, IMF2 is nearest to the canonical alpha band while IMF1 is closest to the canonical beta band. The Residue signal resulting from subtracting IMF2 and IMF1 from the original signal contains mostly delta and some theta peaks as expected.

**Supplementary Figure 3.**
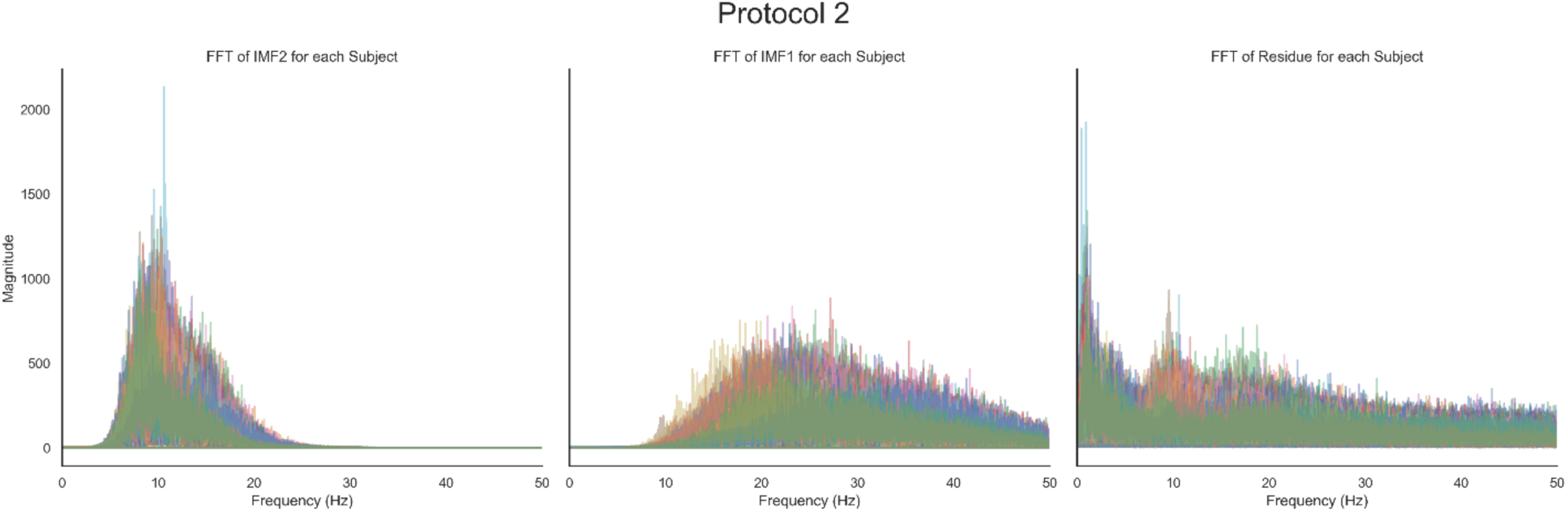
FFT of IMF2, IMF1, and the Residue for Protocol 2 at electrode F3 for each subject. Similar to Protocol 1, from visual inspection, IMF2 is nearest to the canonical alpha band while IMF1 is closest to the canonical beta band. The Residue signal resulting from subtracting IMF2 and IMF1 from the original signal contains mostly delta and some theta peaks as expected.

**Supplementary Table 1.** Prediction Model Comparison Protocol 1. Prediction of treatment outcome specific to Protocol 1 using various methods. For EMD-SBL we use IMF1, IMF2, and the residue signal of the EEG. Note that IMF1 and IMF2 typically was inside the Beta and Alpha frequency bands for this dataset. For SBLEST + Band filtering, we replace IMF1 and IMF2 with the original signal bandpass filtered at 13-40 Hz and 8-13 Hz respectively. Only EMD-SBL achieved a significant (p<0.05) Pearson’s r value.

| Model | PEARSON'S CORRELATION<br>COEFFICIENT (R) | p |
| --- | --- | --- |
| SBLEST | -0.00160 | 0.992 |
| SBLEST w/ Bandpass | 0.0199 | 0.898 |
| Ridge Regression w/ itEMD | 0.166 | 0.280 |
| EMD-SBL | 0.401 | 0.00705 |

**Supplementary Table 2.** Prediction Model Comparison Protocol 2. Prediction of treatment outcome specific to Protocol 2 using various methods. For EMD-SBL we use IMF1, IMF2, and the residue signal of the EEG. Note that IMF1 and IMF2 typically was inside the Beta and Alpha frequency bands for this dataset. For SBLEST + Band filtering, we replace IMF1 and IMF2 with the original signal bandpass filtered at 13-40 Hz and 8-13 Hz respectively. Only EMD-SBL achieved a significant (p<0.05) Pearson’s r value.

| Model | PEARSON'S CORRELATION<br>COEFFICIENT (R) | p |
| --- | --- | --- |
| SBLEST | 0.121 | 0.309 |
| SBLEST w/ Bandpass | 0.151 | 0.201 |
| Ridge Regression w/ itEMD | 0.159 | 0.179 |
| EMD-SBL | 0.255 | 0.0292 |

**Supplementary Table 3.** Prediction Model Comparison Eyes Closed vs Eyes open. Prediction of treatment outcome for both Protocol 1 and Protocol 2 using eyes-open or eyes-closed rsEEG. EMD-SBL with IMF1, IMF2, and the residue signal of the EEG was used for both rsEEG cases. Both modalities were validated using 10 run 10-fold cross validation.

| **rTMS**<br>**Protocol** | **Eyes- Open** | **Eyes- Closed** |
| --- | --- | --- |
| **Protocol 1** | Pearson's r = 0.401, p = 0.00705 | Pearson's r = -0.194, p = 0.235 |
| **Protocol 2** | Pearson's r = 0.255, p = 0.0292 | Pearson's r = 0.188, p = 0.161 |

